# Diagnostic performance of fractional exhaled nitric oxide for asthma in children

**DOI:** 10.64898/2026.04.16.26351005

**Authors:** Mari Sasaki, Myrofora Goutaki, Carmen C.M. de Jong, Pascal Heer, Nicolas Regamey, Alexander Moeller, the SPAC Study Team, Claudia E. Kuehni

## Abstract

**Background:** Recent guidelines differ in how fractional exhaled nitric oxide (FeNO) is used to diagnose school-age asthma, either as one of several tests with a cut-off at 25 ppb or as a single rule-in test at 35 ppb. Evidence on its diagnostic performance and clinical utility in subgroups remain limited.

**Methods:** We analysed data from 1,979 school-age children in the Swiss Paediatric Airway Cohort referred for suspected asthma. We investigated FeNO performance with diagnosis by paediatric pulmonologists as reference standard using receiver operating characteristics curves, selected cut-offs and simulated predictive values across different prevalence. Subgroup analyses considered allergic sensitisation with allergic rhinitis and current inhaled corticosteroid (ICS) use.

**Results:** In the overall cohort (asthma diagnosis 70%), FeNO showed poor discrimination for asthma (AUC 0.66; 95% CI 0.64–0.68) with an optimal cut-off at 22 ppb. At 25 and 35 ppb, sensitivity was low (43%, 95% CI 40–46; 31%, 95% CI 29–34) and specificity moderate to high (84%, 95% CI 77–84; 90%, 95% CI 87–92). Positive predictive value at 35 ppb was 88% and was 57% when simulated at a prevalence of 30%. FeNO had no diagnostic value in non-sensitised children and lower performance in sensitised children with allergic rhinitis than in those without (AUC 0.59 vs 0.68). Current ICS use did not influence performance.

**Conclusion:** FeNO has limited diagnostic performance as a stand-alone test for school-age asthma, and underlying asthma prevalence and allergic characteristics should be considered in the interpretation.

## Introduction

Fractional exhaled nitric oxide (FeNO) has varying roles in clinical guidelines for diagnosing asthma in school-age children. The 2021 European Respiratory Society (ERS) guideline recommends that diagnosis requires at least two positive tests, one of which can be FeNO with a suggested cut-off of 25 ppb (1). In contrast, the 2024 British Thoracic Society (BTS), National Institute for Health and Care Excellence (NICE), and Scottish Intercollegiate Guidelines Network (SIGN) guideline places FeNO at the first step of the algorithm and considers FeNO ≥ 35 ppb sufficient to confirm asthma (2). The 2025 Global Initiative for Asthma guideline update similarly states that FeNO ≥ 35 ppb can support diagnosis when symptoms are typical and lung function is unavailable or normal (3).

These differences reflect limited evidence on the diagnostic performance and clinical utility of FeNO. In the Swiss Paediatric Airway Cohort (SPAC), we previously showed low sensitivity (48%) and moderate specificity (87%) at a cut-off of 23 ppb, consistent with guideline evidence reviews (4–13). However, large clinical studies directly comparing guideline suggested cut-offs are lacking. Furthermore, the clinical impact of using FeNO alone to rule-in asthma, as proposed by the BTS/NICE/SIGN guideline remain unclear. Because the ability of a test to rule in disease, expressed as the positive predictive value (PPV), depends on disease prevalence, the clinical utility of FeNO with this approach may vary across settings with different asthma prevalence (i.e., primary care versus tertiary care). FeNO levels are also affected by allergic sensitisation, allergic rhinitis, and inhaled corticosteroids (ICS) use, which may influence diagnostic performance (14–20). Its clinical utility may therefore differ in situations where information on allergic status or treatment is available and whether these are present. Few studies have evaluated test performance across these population subgroups (6–8, 10).

Previously, we assessed performance of several tests for asthma diagnosis in a smaller sample (n = 514) without exploring guideline-recommended cut-offs or subgroup effects (12). In the present study, we analysed a cohort of nearly 2,000 school-age children referred to paediatric pulmonology clinics in Switzerland for evaluation of possible asthma, enabling more detailed analyses. First, we assessed diagnostic performance and identified the optimal FeNO cut-off for childhood asthma. Second, we evaluated the clinical utility of using FeNO to rule in asthma, as recommended by the BTS/NICE/SIGN guideline across varying asthma prevalence reflecting different clinical setting. Third, we examined how diagnostic performance varies across subgroups defined by allergic sensitisation, allergic rhinitis, and ICS use, to assess its utility when this information is available for clinical decision making.

## Material and methods

### Swiss Paediatric Airway Cohort (SPAC) and study population

SPAC is an ongoing prospective multicentre study that consecutively invites 0–17-year-old children referred to participating paediatric pulmonology clinics for evaluation of suspected asthma or other common respiratory problems (wheeze, chronic cough or exercise-induced symptoms). Children with rare lung diseases (e.g., cystic fibrosis) or with chronic respiratory conditions secondary to neuromuscular disorders or congenital malformations, are not invited to participate. The current analysis includes children newly referred aged 5-17 years to nine participating clinics (Aarau, Basel, Bern, Chur, Horgen, Lausanne, Lucerne, St. Gallen and Zurich). We included all eligible participants with data available in the study database on 14^th^ May 2025.

The procedures of SPAC (clinicaltrials.gov NCT03505216) have been published previously (12, 21). We extract clinical data from the outpatient clinics, including test results, treatments, and diagnoses. Participants complete a standardised questionnaire at the time of the clinical visit and annually about respiratory symptoms and treatments. Ethical approval was granted from the ethics committee of the canton of Bern (2016–02176) and all participating parents and adolescents aged 14 years or older provided written informed consent.

### Asthma diagnosis procedure and the reference standard

Diagnostic procedures at the SPAC clinics followed local protocols and tests were conducted by experienced staff blinded to the diagnosis. Children usually received clinical evaluation, spirometry, FeNO measurement, allergy testing (skin prick test, serum specific IgE or ImmunoCAP Rapid) and bronchodilator reversibility (BDR) testing at the first visit. When indicated, bronchial challenge testing with methacholine, exercise, or other methods was conducted during the first or at a later visit. Paediatric pulmonologists made diagnoses based on all available clinical and test information. For this analysis, we used the diagnosis recorded by the physician as “definite asthma” or “probable asthma” as the reference standard and compared it with children who received a different diagnosis.

### FeNO measurement

FeNO was measured by online single-breath method at a constant flow rate of 50mL/s as the index test. Six clinics used ANALYZER CLD 88 sp (Eco Medics AG, Duernten, Switzerland), and three used Niox MINO or Niox VERO (NIOX Group plc, Oxford, UK), in accordance with published technical standards (22). These devices show good agreement (23).

All clinics measured in doublets before spirometry and reported the mean value of the two measurements.

### Definition of allergic status and inhaled corticosteroid use

We classified children as sensitised if they had evidence of sensitisation to any aeroallergen at the clinical visit or from a previous test, during the past two years. We classified children receiving ICS alone or combined with long-acting β2-agonist (ICS-LABA) as using ICS at the visit. We defined current ICS use based on medical records at the clinical visit, or, when unavailable, from the questionnaire completed at the visit, which asked about treatment in the previous 12 months.

## Statistical analysis

We assessed FeNO performance for asthma diagnosis using receiver operating characteristic (ROC) curves and the area under the curve (AUC), which reflects the ability of the test to discriminate between children with and without asthma across all cut-offs. We interpreted AUC values using conventional thresholds proposed: 0-5-0.7: poor, 0.7–0.8 acceptable, 0.8–0.9 excellent, and > 0.9 outstanding discrimination, recognising that these categories are arbitrary and clinical relevance depends on test use, such as for diagnosis, screening or prediction (24). We tested the AUC against 0.5, which represents no discriminative ability, with DeLong’s method. To assess test performance for specific cut-offs, we calculated sensitivity, specificity, positive and negative predictive values (PPV, NPV), and Youden’s index (sensitivity + specificity – 1) for published, guideline-recommended, and optimal cut-offs that maximised Youden’s index.

Using sensitivity and specificity at selected cut-offs from the first analysis, we simulated and plotted PPV and NPV across varying asthma prevalence to reflect different clinical settings. We selected asthma prevalence of 30%, as an estimate for primary care which is also used in the ERS guideline meta analyses, and 50%, representing the lower end of the 48–72% range reported in predominantly tertiary care studies (1, 2).

We assessed performance across subgroups by stratifying children according to allergic sensitisation and parent-reported allergic rhinitis. Although both are closely related and affect FeNO, sensitisation reflects underlying allergic immune reactivity, whereas allergic rhinitis reflects clinical allergy. Their combination may influence FeNO and diagnostic performance differently than either alone. We compared AUCs between subgroups where the AUC differed from 0.50, using DeLong’s method with Benjamini-Hochberg adjustment. We similarly compared FeNO performance between children with and without ICS use. In a sensitivity analysis, we analysed a subgroup of children with treatment information obtained from the medical records, excluding those with only parent-reported data. The use of physician diagnosis, based on all clinical information including FeNO, as the reference standard may overestimate the diagnostic performance. Therefore, we conducted sensitivity analyses using alternative reference standards: (1) definite asthma versus probable or no asthma, and (2) asthma defined by ICS or ICS-LABA use at one-year follow-up (or two-year if missing) versus no use. We performed complete-case analyses for each variable without missing imputation, using R version 4.4.1 with packages pROC, dplyr and ggplot2 (25–28). The details of the variable definitions and sensitivity analyses are in the online supplementary material.

### Reporting

We followed the Standards for Reporting Diagnostic Accuracy Studies (STARD) check list (29).

## Results

### Study population

Of the 2,262 children aged 5-17 years newly referred to the SPAC clinics, 1,979 (median age 10, IQR 7-12 years) had FeNO measured by online single breath method (Supplementary figure 1). Frequency of FeNO testing varied only slightly between centres (81-94%) and was more than 85% in six out of the nine clinics (Supplementary table 1). Children without online single breath FeNO measurements were younger, reported wheeze triggered by exercise less often, and had more emergency visits or hospitalisations for respiratory symptoms than those with measurements (Supplementary table 2).

Paediatric pulmonologists diagnosed asthma in 70% of the 1,979 children: 969 (49%) with definite asthma and 412 (21%) with probable asthma. Children not diagnosed as having asthma received a range of diagnoses, including extra-thoracic or thoracic dysfunctional breathing, allergic rhinitis or allergy related respiratory symptoms and cough of unknown aetiology (Supplementary table 3). Children diagnosed with asthma were slightly younger (10 vs 11 years, Table 1), more often boys (59% vs 49%) and more frequently reported respiratory symptoms, including wheeze (84% vs 52%), and allergic rhinitis (45% vs 32%) than those without asthma. Proportions of children with allergic sensitisation (86% vs 64%) and with ICS or ICS-LABA use (50 vs 24%) were also higher in children diagnosed with asthma.

**Table 1.** Clinical characteristics of all school-age children under investigation of asthma in the Swiss Paediatric Airway Cohort comparing children with and without asthma diagnosis.

| Characteristics | Overall<br>N = 1,979 | Diagnosis<br>Asthma<br>N = 1,381 | Not asthma<br>N = 598 | p-value |
| --- | --- | --- | --- | --- |
| Age in years, median (IQR) | 10.0 (7.0, 12.0) | 10.0 (7.0, 12.0) | 11.0 (8.0, 13.0) | <0.001 |
| Male | 1,105 (56%) | 812 (59%) | 292 (49%) | <0.001 |
| BMI z-score, median (IQR) | 0.22 (-0.43, 1.02) | 0.23 (-0.44, 1.00) | 0.19 (-0.40, 1.08) | 0.96 |
| Self-reported symptoms and events in the last 12 months |  |  |  |  |
| Any wheeze | 1,469 (74%) | 1,156 (84%) | 313 (52%) | <0.001 |
| More than three attacks | 514 (26%) | 431 (31%) | 83 (14%) | <0.001 |
| Wheeze triggered by cold | 1,040 (53%) | 859 (62%) | 181 (30%) | <0.001 |
| Wheeze triggered by exercise | 1,141 (58%) | 897 (65%) | 244 (41%) | <0.001 |
| Wheeze triggered by pollen | 527 (27%) | 461 (33%) | 66 (11%) | <0.001 |
| Wheeze triggered by dust | 282 (14%) | 256 (19%) | 26 (4.3%) | <0.001 |
| Wheeze triggered by pets | 242 (12%) | 225 (16%) | 17 (2.8%) | <0.001 |
| Awakening due to wheeze | 597 (30%) | 519 (38%) | 78 (13%) | <0.001 |
| Dyspnoea due to wheeze | 855 (43%) | 737 (53%) | 118 (20%) | <0.001 |
| Emergency visit due to respiratory symptoms | 729 (37%) | 572 (41%) | 157 (26%) | <0.001 |
| Hospitalisations due to respiratory symptoms | 153 (7.7%) | 144 (10%) | 9 (1.5%) | <0.001 |
| Cough more than others | 945 (48%) | 702 (51%) | 243 (41%) | <0.001 |
| Self-reported allergic rhinitis | 810 (41%) | 618 (45%) | 192 (32%) | <0.001 |
| Allergic sensitisation (n = 1,457)<sup>#</sup> | 1,153 (79%) | 879 (86%) | 274 (64%) | <0.001 |
| Prior ICS-LABA or ICS use (n = 1,764)<sup>#</sup> | 740 (42%) | 607 (50%) | 133 (24%) | <0.001 |
| FeNO geometric mean, 95%CI (ppb) | 17.7 (17.0-18.5) | 20.6 (19.6-21.7) | 12.5 (11.7-13.3) | <0.001 |

Abbreviations, FeNO: fractional exhaled nitric oxide, ppb: parts per billion, IQR: interquartile range, BMI: body mass index, ICS-LABA: inhaled corticosteroid and long-acting $\beta$ -agonist, ICS: inhaled corticosteroid
p-value was derived by Pearson's Chi-squared test or Wilcoxon rank sum test.
<sup>#</sup>Information on allergic sensitisation and ICS-LABA/ICS use were available in 1,435 and 1,764 out of the 1,979 children in the analysis.

### Diagnostic performance of FeNO among children in SPAC

FeNO showed poor discrimination (AUC) for asthma at 0.66 (95%CI: 0.63-0.68, p-value: < 0.001, Figure 1) and the optimal cut-off by Youden’s index was 22 ppb, with a sensitivity of 48% (95%CI: 45-50) and a specificity of 81% (95%CI: 77-84). At 25 ppb (ERS-suggested cut-off), and 35 ppb (BTS/NICE/SIGN-suggested cut-off), sensitivity was 43% (95% CI 40–46) and 31% (95% CI 29–34), specificity 84% (95% CI 81–87) and 90% (95% CI 87–92), PPV 86% (95% CI 83–89) and 88% (95% CI 85–91), and NPV 39% (95% CI 36–42) and 36% (95% CI 34–39). Overall, diagnostic parameters at 22 and 25 ppb were similar. A sensitivity of 95% would only be achieved with a cut-off of 4.5 ppb, which would however result in a specificity of 6%, whereas 95% specificity requires a cut-off of 44 ppb and results in a sensitivity of 24% (Supplementary table 4).

**Figure 1.**
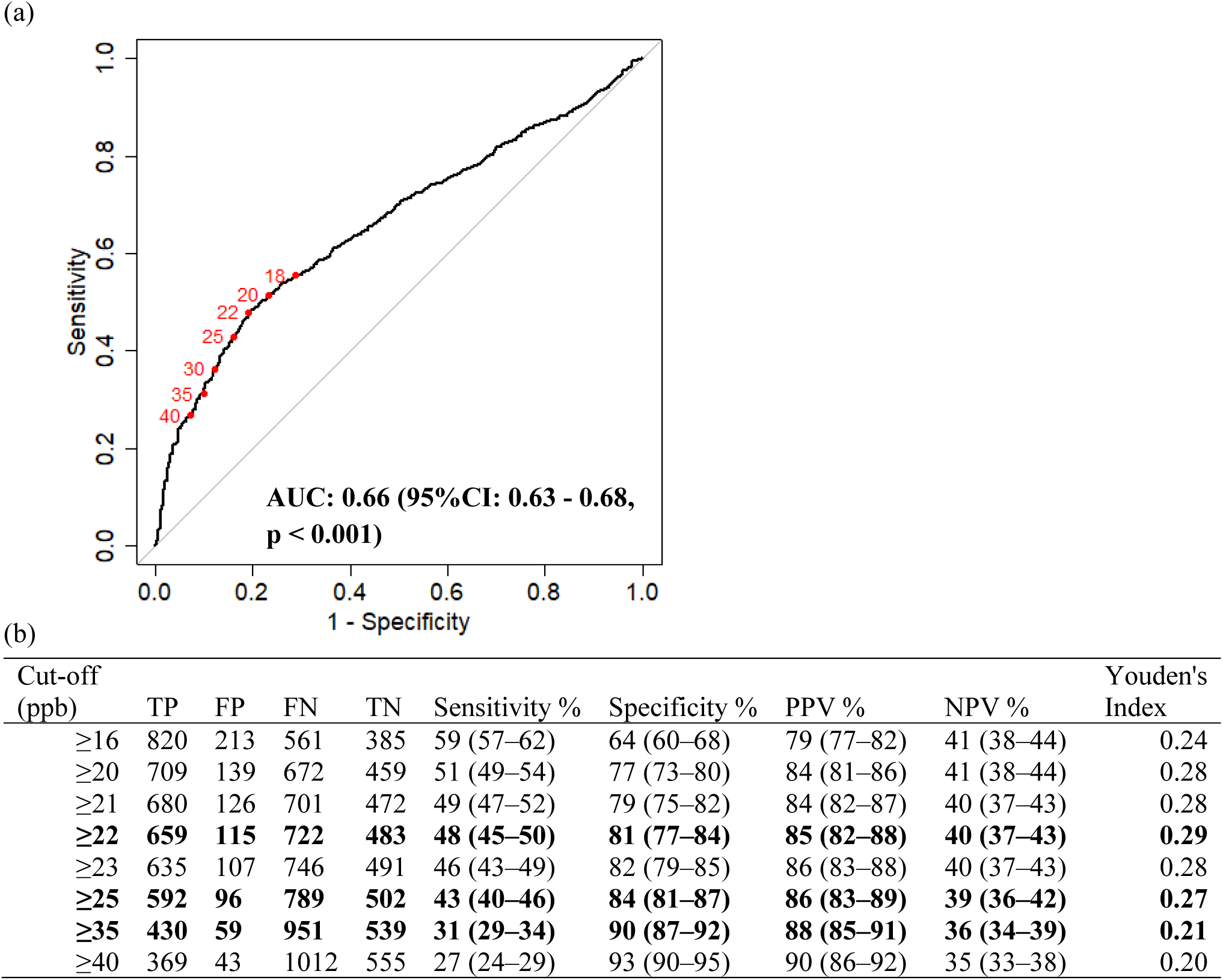
Diagnostic performance of fractional exhaled nitric oxide (FeNO) for asthma among school-age children in the Swiss Paediatric Airway Cohort (n = 1,979, asthma 70%) (a) Receiver operating characteristic (ROC) curve for FeNO, values in red denote the cut-off points corresponding to different sensitivity/specificity combinations. P-value is derived by testing the AUC against 0.5 using DeLong’s method. (b) Diagnostic performance at selected FeNO cut-off values. Data are presented as n or n (95%CI), values with the cut-off for the highest Youden’s index (22 ppb), ERS cut-off (25 ppb) and BTS/NICE/SIGN cut-off (35 ppb) are presented in bold. Abbreviations, AUC: area under the curve, TP: true positive, FP: false positive, FN: false negative, TN: true negative, PPV: positive predictive value, NPV: negative predictive value, FeNO: fractional exhaled nitric oxide, ppb: parts per billion

### Clinical utility of using FeNO to rule-in asthma according to BTS/NICE/SIGN guideline

Considering a FeNO cut-off of 35 ppb to rule-in asthma as proposed in the BTS/NICE/SIGN guideline in our study, 59 (12%) of the 489 children with FeNO ≥ 35 ppb, did not receive an asthma diagnosis (i.e., false positive cases). Children without asthma most commonly had allergic rhinitis or allergy-related respiratory symptoms (37%) as the main diagnosis (Table 2). Using the sensitivity and specificity observed at the 22, 25 and 35 ppb cut-offs in the first analysis, simulated PPVs were similar across the three cut-offs for each prevalence (Figure 2). In these simulations, the estimated PPV for the 35-ppb cut-off was above 80% when asthma prevalence was higher than around 60%. However, at lower prevalence of 30%, reflecting non-tertiary or primary care settings, simulated PPV was lower at 57%, corresponding to false positive rate of 43%.

**Figure 2.**
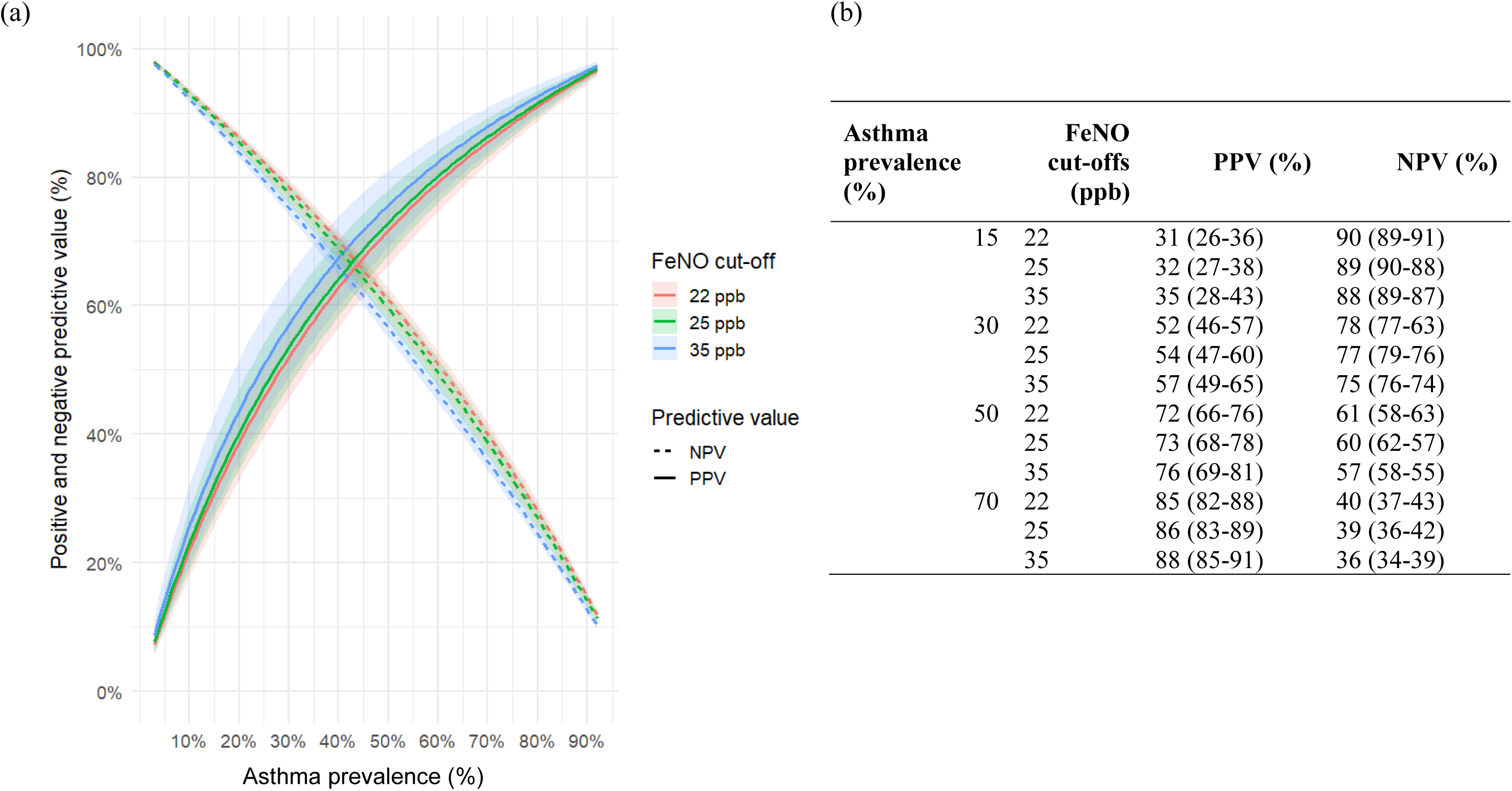
Simulated positive and negative predictive values of fractional exhaled nitric oxide (FeNO) for asthma across different asthma prevalences in the Swiss Paediatric Airway Cohort (a) PPV and NPV across a range of prevalences based on the observed diagnostic performance. Solid line = probability of asthma if FeNO ≥ cut-off (PPV); dashed line = probability of not having asthma if FeNO < cut-off (NPV). For example, at a prevalence (pre-test probability) of 15%, FeNO ≥35 ppb gives a 35% probability of asthma, while FeNO <35 ppb gives 88% probability of not having asthma. Shaded ribbons indicate 95% confidence intervals derived from the sensitivity and specificity estimates. (b) Selected PPV and NPV values at selected prevalence levels. Abbreviations PPV: positive predictive value, NPV: negative predictive value, FeNO: fractional exhaled nitric oxide, ppb: parts per billion

**Table 2.** Diagnoses among school-age children potentially over-diagnosed using the fractional exhaled nitric oxide (FeNO) ≥ 35 ppb rule-in approach (BTS/NICE/SIGN) in the Swiss Paediatric Airway Cohort.

| **Final diagnosis** | **Not asthma**<br>**N = 59** |
| --- | --- |
| Allergic rhinitis or allergy related respiratory symptoms | 22 (37%) |
| Upper airway cough | 8 (14%) |
| Bronchial hyper-responsiveness (no clinical asthma) | 7 (12%) |
| Exercise induced symptoms of unknown aetiology | 8 (14%) |
| Extra-thoracic dysfunctional breathing | 5 (9%) |
| Thoracic dysfunctional breathing | 4 (7%) |
| Other<sup>#</sup> | 5 (9%) |

<sup>#</sup> Other diagnoses include chronic cough of unknown aetiology, habitual cough and upper airway issues.

### Clinical utility of FeNO in relevant subgroups

#### Based on allergic sensitisation and allergic rhinitis

In the two subgroups of non-sensitised children with and without report of allergic rhinitis, AUCs did not differ from 0.5 (both p-adj = 0.58) indicating no discrimination for asthma by FeNO (Figure 3). In sensitised children, FeNO discriminated asthma in both subgroups with and without allergic rhinitis, but poorly (AUC <0.7). There, AUC was slightly higher in those without allergic rhinitis compared to those without (AUC, 95%CI: 0.68, 0.63-0.72 vs 0.59, 0.54-0.65, p = 0.02). Optimal cut-offs were similar to the overall cohort at 20 to 22 ppb in sensitised children with and without allergic rhinitis. Between sensitised children with and without allergic rhinitis, sensitivity was similar at the 22, 25 and 35 ppb cut-offs. Specificity was lower in children with allergic rhinitis (e.g., 78% vs 88% at 35 ppb), although 95%CIs partly overlapped.

**Figure 3.**
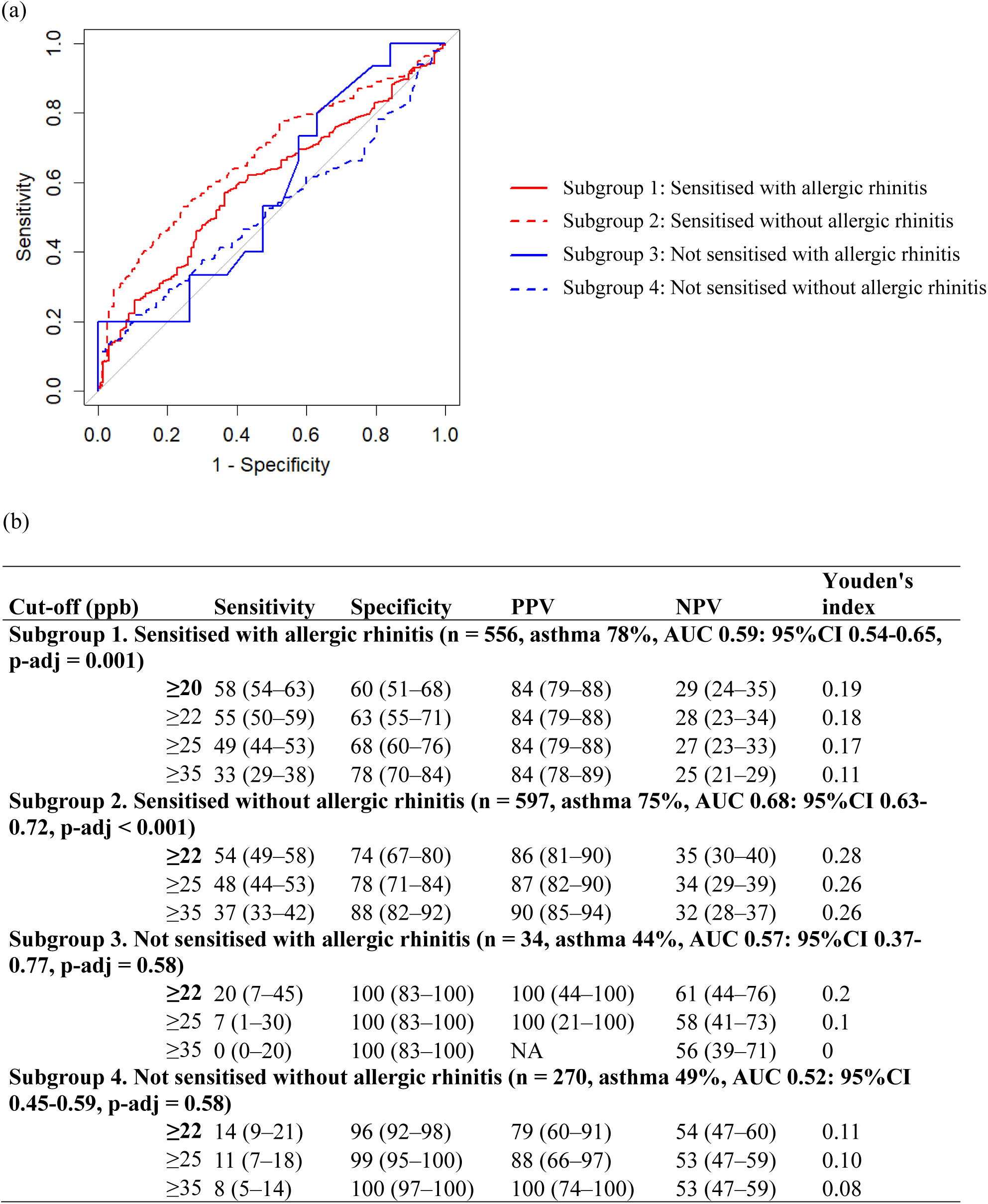
Subgroup analysis of the diagnostic performance of fractional exhaled nitric oxide (FeNO) for asthma in the Swiss Paediatric Airway Cohort by allergic sensitisation and allergic rhinitis (a) Receiver operating characteristic (ROC) curve for FeNO in each subgroup by the combination of allergic sensitisation and allergic rhinitis. (b) Diagnostic performance in each subgroup at selected FeNO cut-off values. Data are presented as n or n (95%CI), values with the cut-off with the highest Youden’s index within each subgroup (bold), optimal cut-off among all children in SPAC (22 ppb), ERS cut-off (25 ppb) and BTS/NICE/SIGN cut-off (35 ppb). NA for PPV in Subgroup 3 is due to no children with FeNO ≥ 35 ppb in this subgroup. Adjusted p-values were obtained by testing each AUC against 0.5 using DeLong’s method and controlling the false discovery rate (Benjamini–Hochberg correction). DeLong’s test for AUC difference between Subgroup 1 and Subgroup 2: p = 0.02. Abbreviations AUC: area under the curve, PPV: positive predictive value, NPV: negative predictive value, FeNO: fractional exhaled nitric oxide, ppb: parts per billion

### Based on ICS use

AUCs for FeNO were similar between children with and without ICS use (AUC, 95%CI: 0.68, 0.64-0.73 vs 0.67, 0.64-0.71, respectively, p = 0.73) and were comparable to the overall cohort (Figure 4). The optimal cut-off was at 21 and 22 ppb in the two subgroups. At the 22, 25 and 35 ppb cut-offs, the differences in sensitivity and specificity were small with overlapping 95%CIs. Results were similar in the subgroup of 448 children who had available treatment information from the medical records (Supplementary figure 2).

**Figure 4.**
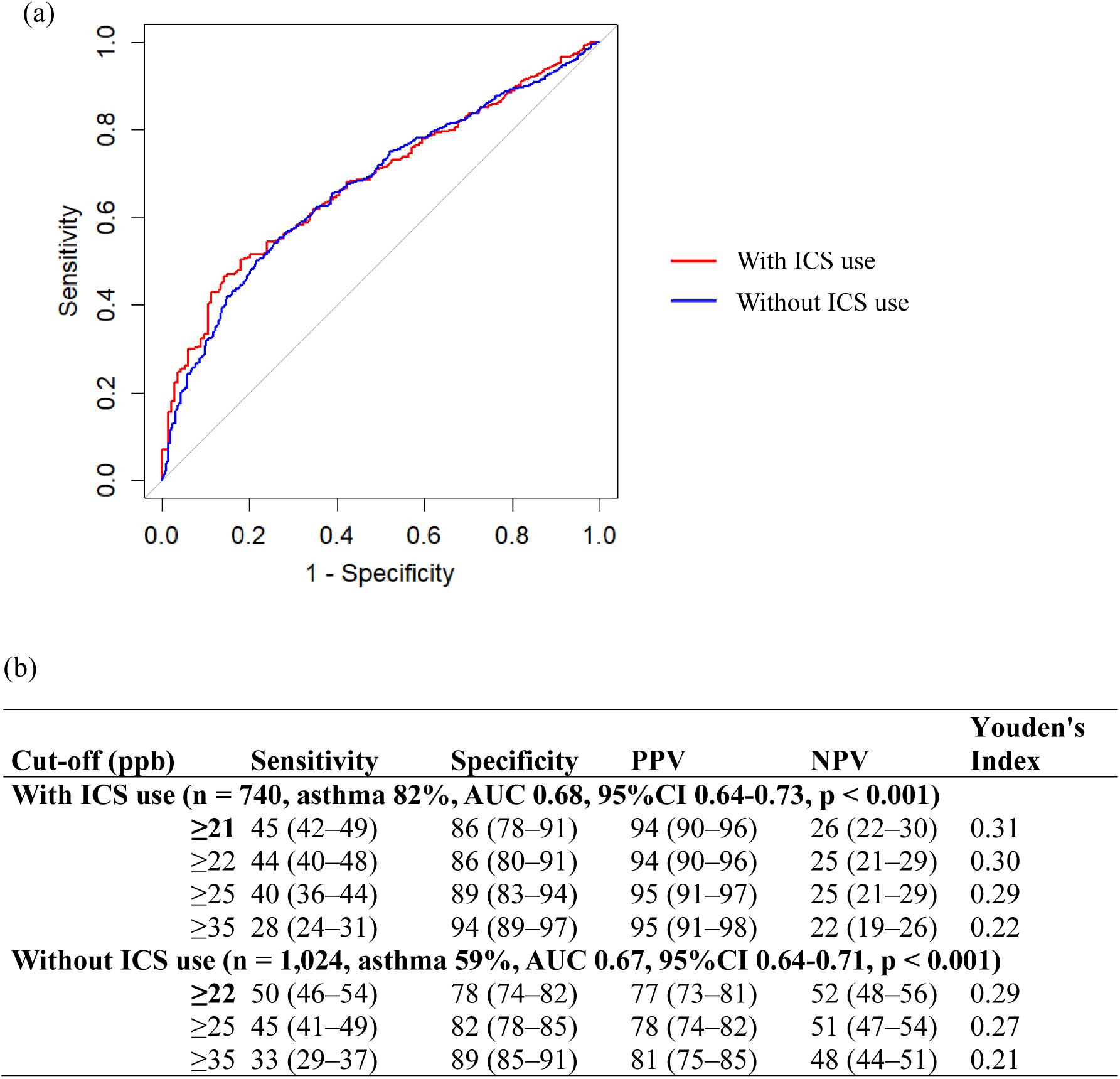
Subgroup analysis of the diagnostic performance of fractional exhaled nitric oxide (FeNO) for asthma in the Swiss Paediatric Airway Cohort by current inhaled corticosteroid use (a) Receiver operating characteristic (ROC) curve for FeNO in children with and without ICS use. (b) Data are presented as n or n (95%CI), values with the cut-off with the highest Youden’s index within each subgroup (bold), optimal cut-off among all children in SPAC (22 ppb), ERS cut-off (25 ppb) and BTS/NICE/SIGN cut-off (35 ppb). P- values were obtained by testing each AUC against 0.5 using DeLong’s method. DeLong’s test for AUC difference between with vs without ICS use: p = 0.73. Abbreviations, AUC: area under the curve, ICS: inhaled cortico-steroid, PPV: positive predictive value, NPV: negative predictive value, FeNO: fractional exhaled nitric oxide, ppb: parts per billion

### Sensitivity analysis using different reference standards

In the sensitivity analyses using alternative reference standards, AUC of the overall cohort was unchanged from the primary analysis when combining the probable asthma to the group without asthma as controls (AUC 0.66, 95%CI 0.63-0.68, p < 0.001, Supplementary figure 3).

AUC was lower when using ICS at 1-2 years as an alternative outcome of asthma (AUC 0.58, 95%CI 0.55-0.62, p < 0.001).

## Discussion

This pragmatic study of a large cohort of school-age children referred for suspected asthma shows that FeNO as a single diagnostic test has poor discrimination, with low sensitivity and moderate to high specificity at optimal and guideline-recommended cut-offs. The clinical utility of FeNO to rule in asthma (PPV) at 35 ppb was high in this cohort of children seen by specialists but simulated PPV decreased substantially in lower prevalence, reflecting primary care. FeNO has no diagnostic value in children without allergic sensitisation and shows lower discrimination and specificity in sensitised children with allergic rhinitis. Diagnostic performance did not differ based on current ICS use.

The overall diagnostic performance of FeNO aligns with previous studies. Papers in the ERS and BTS/NICE/SIGN evidence reviews reported AUCs ranging from 0.53 to 0.80 in most studies and reaching 0.90 in only two (5, 6, 8–11, 13, 30). In the ERS review, mean sensitivity and specificity across five studies were 0.57 and 0.81 at 25 ppb (1, 4, 6, 9, 10, 13). The BTS/NICE/SIGN review similarly reported limited sensitivity, reaching only 0.60 at > 20 ppb in high-quality evidence, while most studies reported specificities above 0.80 (2, 5, 7, 8, 10, 11, 30).

Findings on subgroup influences on FeNO performance partly agreed with previous studies. Several studies have similarly reported no diagnostic value in non-sensitised children or increased FeNO levels in children with allergic sensitisation or rhinitis independent of asthma (6, 8, 10). However, evidence on the combined influence of allergic sensitisation and rhinitis on performance remains limited. In contrast to our results, a Polish study reported that FeNO was associated with asthma among sensitised children with allergic rhinitis but not without allergic rhinitis (7). Higher prevalence of allergic rhinitis in that study (51%) compared to SPAC (41%), or its definition being clinician diagnosis (vs self-report in SPAC) may explain this inconsistency. Evidence on the effect of ICS use on FeNO performance is also limited. Recent guideline reviews did not examine this, and a previous review comparing overall and steroid-naïve populations across mixed age groups and study designs reported inconclusive results (1, 2, 31). In our study, the absence of an effect may reflect that prescribed ICS were not consistently used, or it was insufficient to suppress airway inflammation at the time of assessment. (32, 33).

The limited diagnostic performance of FeNO supports guideline recommendations that it should not be used as the only diagnostic test alongside other tests (1, 2). Sensitivity and specificity were best balanced at 22 ppb and close to the ERS cut-off. Conversely, applying a 35 ppb rule-in approach, as proposed in the BTS/NICE/SIGN algorithm may lead to substantial false positives in lower asthma prevalence settings (2). FeNO is not currently used in Swiss primary care but its use may increase with recent guideline recommendations, although access remains limited in the United Kingdom despite guidelines and national initiatives (34, 35). Further evaluation of FeNO within full diagnostic algorithms and in primary or non-tertiary settings is needed.

Our results suggest that the clinical utility of FeNO differs when allergic status is known and incorporated into diagnostic decision making. In children with suspected asthma who are known not to be sensitised, FeNO adds no diagnostic value. In children with allergic sensitisation, FeNO discriminates asthma less well and is less specific when allergic rhinitis is present, reducing its utility. Although the difference in specificity at 25 or 35-ppb was modest and confidence intervals partly overlapped, it may affect interpretation when FeNO values are close to these cut-offs. Diagnostic performance was similar in children with and without ICS use, suggesting that FeNO may be interpreted without adjustment for prior ICS treatment, but this should be evaluated further.

The key strength of this study is the large multicentre clinical cohort of children investigated for suspected asthma, allowing detailed subgroup analyses. Variation in clinical assessment between centres reflects real world practice and supports generalisability of our findings. Additionally, FeNO was measured in most children referred to these clinics. Children with measurements were older, likely explaining their ability to complete the test and the observed clinical differences. This is consistent with routine clinical practice and is unlikely to introduce substantial selection bias. The main limitation is the use of physician diagnosis as the reference standard, based on all tests including FeNO, which may overestimate its performance (36). However, physician diagnosis reflects routine clinical practice and remains common in the absence of a universally accepted reference standard (37, 38). Despite this potential over estimation, FeNO showed limited performance, strengthening our findings. Sensitivity analyses using alternative reference standards showed similar findings and did not change the overall interpretation. Finally, differences in how ICS use was recorded in medical charts and measurement error in parental reports may have led to misclassification of current ICS use and the observed lack of effect on FeNO performance. Our analysis did not aim to assess the biological effects of ICS on FeNO levels. In conclusion, FeNO should be used with caution as a diagnostic test for asthma in school-age children, given its limited performance. Its role as a rule-in test, as proposed in some guidelines, may not perform consistently across settings. Variation in performance by allergic characteristics should be considered when interpreting FeNO in clinical practice and in future guideline recommendations.

## Acknowledgements

The authors would like to thank the children and families who took part in the SPAC study, the research assistants (S. Chellakudam, L. Mangalath, E. Schneider, and L. Volkart) for their contribution to data collection and entry, PedNet Bern for supporting data collection in Bern, and the members of the SPAC Study Team. Members of the current and past SPAC Study Team are: C. Ardura-Garcia (Institute of Social and Preventive Medicine); J. Barben (Children‘s Hospital of Eastern Switzerland); S. Beck (University Children’s Hospital Zurich); L. Benz (University Children’s Hospital Zurich); D. Berger (Institute of Social and Preventive Medicine); C. Bernold (Children’s Hospital of Central Switzerland); C. Bieli (Cantonal Hospital Aarau); S. Blanchon (Lausanne University Hospital); M. Bullo (Inselspital); G. Buggle (University Children’s Hospital Zurich); C. Casaulta (Inselspital); C.C.M. de Jong (Inselspital); C. Fernandez-Elviro (Lausanne University Hospital); B. Frauchiger (Inselspital); S. Glick (Institute of Social and Preventive Medicine); M. Goutaki (Institute of Social and Preventive Medicine); S. Guerin (Lausanne University Hospital); B. Guerra (Institute of Social and Preventive Medicine); P. Heer (Children’s Hospital of Central Switzerland); M. Hitzler (Children’s Hospital of Central Switzerland); K. Hoyler (Private paediatric pulmonologist); K. Hrup (Children’s Hospital of Central Switzerland); P. Iseli (Cantonal Hospital Graubünden); A. Jochmann (University Children’s Hospital Basel); A. Jung (Children‘s Hospital of Eastern Switzerland); E. Kieninger (Inselspital); I. Korten (Inselspital); T. Krasnova (Institute of Social and Preventive Medicine); L. Krüger (Private paediatric practice); C.E. Kuehni (Institute of Social and Preventive Medicine); Y.T. Lam (University Children’s Hospital Zurich); P. Latzin (Inselspital); M. Lurà (Children’s Hospital of Central Switzerland); R. Makhoul (Institute of Social and Preventive Medicine); M.C. Mallet (Institute of Social and Preventive Medicine); A. Moeller (University Children’s Hospital Zurich); E. Pedersen (Institute of Social and Preventive Medicine); N. Regamey (Children’s Hospital of Central Switzerland); I. Rochat (Lausanne University Hospital); F. Romero (Institute of Social and Preventive Medicine); M. Sasaki (Institute of Social and Preventive Medicine); T. Schürmann (Cantonal Hospital Aarau); E. Seidl (University Children’s Hospital Zurich); E. Sidler (Children’s Hospital of Central Switzerland); G. Signorelli (University Children’s Hospital Zurich); O. Sutter (Private paediatric practice); D. Trachsel (University Children’s Hospital Basel); J. Usemann (University Children’s Hospital Basel); B. Vomsattel (Inselspital); V. Weihrauch (Institute of Social and Preventive Medicine); S. Yammine (Inselspital).

## Support statement

This study was funded by the Swiss National Science Foundation (SNF 212519).

## Author contributions

CK and MG conceptualised the study. CdeJ, AM, PH and NR recruited patients and took part in the investigation. MS conducted the analysis. CK and MG supervised the research. MS drafted the manuscript, and all authors contributed to its revision. All authors approved the final version of the manuscript.

## Conflict of interest

MS, MG, CdeJ, PH and CK have nothing to disclose. AM reports personal fees and grants from Vertex—all outside the submitted work. NR reports personal fees from OM Pharma, Schwabe Pharma, Vertex, and Sanofi—all outside the submitted work.

## Data availability statement

SPAC study data is not deposited at an open access repository, as participants were not asked to give consent to have their data deposited publicly. Requests for partial datasets for specific analyses including individual patient data and a data dictionary defining each included field can be addressed to Prof Kuehni upon reasonable request.

## Notes

### Author Declarations

The Bern Cantonal Ethics Committee (Kantonale Ethikkommission Bern 2016-02176) approved the study; written informed consent was obtained from parents and patients aged ≥ 14 years.

## References

1. Gaillard EA, Kuehni CE, Turner S, Goutaki M, Holden KA, de Jong CCM, et al. European Respiratory Society clinical practice guidelines for the diagnosis of asthma in children aged 5-16 years. Eur Respir J. 2021;58(5).

2. BTS/NICE/SIGN. Asthma: diagnosis, monitoring and chronic asthma management (BTS, NICE, SIGN), Published: 27 November 2024 [Available from: https://www.nice.org.uk/guidance/indevelopment/gid-ng10186/documents.

3. Global Intiative for Asthma. Asthma management and prevention for adults, adolescents and children 6-11 years (2025). A summary guide for healthcare providers. [updated June 2025. Available from: ginasthma.org.

4. Brouwer AF, Visser CA, Duiverman EJ, Roorda RJ, Brand PL. Is home spirometry useful in diagnosing asthma in children with nonspecific respiratory symptoms? Pediatr Pulmonol. 2010;45(4):326–32.

5. Eom SY, Lee JK, Lee YJ, Hahn YS. Combining spirometry and fractional exhaled nitric oxide improves diagnostic accuracy for childhood asthma. Clin Respir J. 2020;14(1):21–8.

6. Grzelewski T, Witkowski K, Makandjou-Ola E, Grzelewska A, Majak P, Jerzynska J, et al. Diagnostic value of lung function parameters and FeNO for asthma in schoolchildren in large, real-life population. Pediatr Pulmonol. 2014;49(7):632–40.

7. Jerzynska J, Majak P, Janas A, Stelmach R, Stelmach W, Smejda K, et al. Predictive value of fractional nitric oxide in asthma diagnosis-subgroup analyses. Nitric Oxide. 2014;40:87–91.

8. Kessler A, Kragl U, Glass A, Schmidt S, Fischer DC, Ballmann M. Exhaled nitric oxide can’t replace the methacholine challenge in suspected pediatric asthma. Respir Med. 2019;157:21–5.

9. Sivan Y, Gadish T, Fireman E, Soferman R. The use of exhaled nitric oxide in the diagnosis of asthma in school children. J Pediatr. 2009;155(2):211–6.

10. Woo SI, Lee JH, Kim H, Kang JW, Sun YH, Hahn YS. Utility of fractional exhaled nitric oxide (F(E)NO) measurements in diagnosing asthma. Respir Med. 2012;106(8):1103–9.

11. Zhou J, Zhao X, Zhang X, Yu X, Wang Y, Jiang W, et al. Values of fractional exhaled nitric oxide for cough-variant asthma in children with chronic cough. J Thorac Dis. 2018;10(12):6616–23.

12. de Jong CCM, Pedersen ESL, Mozun R, Müller-Suter D, Jochmann A, Singer F, et al. Diagnosis of asthma in children: findings from the Swiss Paediatric Airway Cohort. Eur Respir J. 2020;56(5).

13. de Jong CCM, Pedersen ESL, Mozun R, Goutaki M, Trachsel D, Barben J, et al. Diagnosis of asthma in children: the contribution of a detailed history and test results. Eur Respir J. 2019;54(6).

14. Sunde RB, Thorsen J, Skov F, Hesselberg L, Kyvsgaard J, Folsgaard NV, et al. Exhaled nitric oxide is only an asthma-relevant biomarker among children with allergic sensitization. Pediatr Allergy Immunol. 2023;34(11):e14044.

15. Henriksen AH, Sue-Chu M, Holmen TL, Langhammer A, Bjermer L. Exhaled and nasal NO levels in allergic rhinitis: relation to sensitization, pollen season and bronchial hyperresponsiveness. Eur Respir J. 1999;13(2):301–6.

16. Wang R, Fowler SJ, Turner SW, Drake S, Healy L, Lowe L, et al. Defining the normal range of fractional exhaled nitric oxide in children: one size does not fit all. ERJ Open Res. 2022;8(3).

17. Khatri SB, Iaccarino JM, Barochia A, Soghier I, Akuthota P, Brady A, et al. Use of Fractional Exhaled Nitric Oxide to Guide the Treatment of Asthma: An Official American Thoracic Society Clinical Practice Guideline. Am J Respir Crit Care Med. 2021;204(10):e97–e109.

18. Flashner BM, Rifas-Shiman SL, Oken E, Camargo CA, Jr., Platts-Mills TAE, Workman L, et al. Contributions of asthma, rhinitis and IgE to exhaled nitric oxide in adolescents. ERJ Open Res. 2021;7(2).

19. Krantz C, Accordini S, Alving K, Corsico AG, Demoly P, Ferreira DS, et al. Cross-sectional study on exhaled nitric oxide in relation to upper airway inflammatory disorders with regard to asthma and perennial sensitization. Clin Exp Allergy. 2022;52(2):297–311.

20. Saranz RJ, Lozano A, Lozano NA, Ponzio MF, Cruz AA. Subclinical lower airways correlates of chronic allergic and non-allergic rhinitis. Clin Exp Allergy. 2017;47(8):988–97.

21. Pedersen ESL, de Jong CCM, Ardura-Garcia C, Barben J, Casaulta C, Frey U, et al. The Swiss Paediatric Airway Cohort (SPAC). ERJ Open Res. 2018;4(4).

22. American Thoracic S, European Respiratory S. ATS/ERS recommendations for standardized procedures for the online and offline measurement of exhaled lower respiratory nitric oxide and nasal nitric oxide, 2005. Am J Respir Crit Care Med. 2005;171(8):912–30.

23. Schiller B, Hammer J, Barben J, Trachsel D. Comparability of a hand-held nitric oxide analyser with online and offline chemiluminescence-based nitric oxide measurement. Pediatr Allergy Immunol. 2009;20(7):679–85.

24. Hosmer DW, Jr., Lemeshow, S. Sturdivant, R.X. Applied Logistic Regression: John Wiley & Sons, Inc.; 2013 2013/03/22. 153-225 p.

25. Robin X, Turck N, Hainard A, Tiberti N, Lisacek F, Sanchez J-C, et al. pROC: an open-source package for R and S+ to analyze and compare ROC curves. BMC Bioinformatics. 2011;12:77.

26. Wickham H, ggplot2: Elegant Graphics for Data Analysis: Springer-Verlag New York; 2016 [Available from: https://ggplot2.tidyverse.org.

27. Wickham H, François R, Henry L, Müller K, Vaughan D, dplyr: A Grammar of Data Manipulation 2025 [Available from: https://CRAN.R-project.org/package=dplyr.

28. R Core Team, R: A Language and Environment for Statistical Computing Vienna, Austria 2024 [Available from: https://www.R-project.org/.

29. Bossuyt PM, Reitsma JB, Bruns DE, Gatsonis CA, Glasziou PP, Irwig L, et al. STARD 2015: an updated list of essential items for reporting diagnostic accuracy studies. BMJ. 2015;351:h5527.

30. Livnat G, Yoseph RB, Nir V, Hakim F, Yigla M, Bentur L. Evaluation of high-sensitivity serum CRP levels compared to markers of airway inflammation and allergy as predictors of methacholine bronchial hyper-responsiveness in children. Lung. 2015;193(1):39–45.

31. Wang Z, Pianosi PT, Keogh KA, Zaiem F, Alsawas M, Alahdab F, et al. The Diagnostic Accuracy of Fractional Exhaled Nitric Oxide Testing in Asthma: A Systematic Review and Meta-analyses. Mayo Clinic Proceedings. 2018;93(2):191–8.

32. Menzies-Gow A, Mansur AH, Brightling CE. Clinical utility of fractional exhaled nitric oxide in severe asthma management. Eur Respir J. 2020;55(3).

33. Smith AD, Cowan JO, Brassett KP, Filsell S, McLachlan C, Monti-Sheehan G, et al. Exhaled nitric oxide: a predictor of steroid response. Am J Respir Crit Care Med. 2005;172(4):453–9.

34. AHSN Network. National FeNO programme Impact report 2023 [Available from: https://www.pcrs-uk.org/resource/current/feno-national-programme-impact-report.

35. Asthma + Lung UK. NG245 One Year On: Implementing the BTS/NICE/SIGN asthma guideline [Available from: https://www.asthmaandlung.org.uk/ng245-one-year.

36. Whiting P, Rutjes AW, Reitsma JB, Glas AS, Bossuyt PM, Kleijnen J. Sources of variation and bias in studies of diagnostic accuracy: a systematic review. Ann Intern Med. 2004;140(3):189–202.

37. Umemneku Chikere CM, Wilson K, Graziadio S, Vale L, Allen AJ. Diagnostic test evaluation methodology: A systematic review of methods employed to evaluate diagnostic tests in the absence of gold standard - An update. PLoS One. 2019;14(10):e0223832.

38. Rutjes AW, Reitsma JB, Coomarasamy A, Khan KS, Bossuyt PM. Evaluation of diagnostic tests when there is no gold standard. A review of methods. Health Technol Assess. 2007;11(50):iii, ix-51.

