## Supplementary material for "Diagnostic performance of fractional exhaled nitric oxide for asthma in children"

Supplementary methods

*Definition of variables*

The following variables were defined using responses to the questionnaire items:

- Cough more than others: “Do you think your child coughs more often than other children?”

- Emergency visits due to respiratory symptoms: “Has your child had an unplanned visit to a paediatrician or a hospital emergency department in the past 12 months for cough, wheeze, or asthma?”

- Hospitalisation due to respiratory symptoms: “Has your child been hospitalised in the past 12 months for cough, wheeze, or asthma?”

- Self-reported allergic rhinitis: “Has your child ever had hay fever?”

Body mass index z-scores were calculated based on the WHO 2007 References for School-age Children and Adolescents (5 to 19 years) (1).

*Sensitivity analysis*

The reference standard in the main analysis was the physician diagnosis of asthma (definite or probable asthma) recorded at the clinical visit. This diagnosis was made by the responsible paediatric pulmonologist, based on all available clinical information, including results of fractional exhaled nitric oxide (FeNO) and other diagnostic tests. Because FeNO formed part of the information available to the physician when establishing the diagnosis, the use of this reference standard could overestimate the diagnostic performance of FeNO (2).

To assess the robustness of the findings, we conducted sensitivity analyses using alternative reference standards. First, we compared children classified as having definite asthma with those classified as probable asthma or no asthma, providing a stricter diagnostic definition. Second, we used treatment at follow-up as an alternative indicator of asthma, defining asthma as the use of inhaled corticosteroids (ICS) or ICS combined with long-acting β2-agonist (ICS-LABA) at one-year follow-up (or two-year follow-up if one-year data were missing). We considered ongoing controller treatment a pragmatic proxy for persistent asthma in routine clinical practice. These analyses allowed us to evaluate whether the estimated diagnostic accuracy of FeNO was consistent when different definitions of asthma were applied.

Supplementary figure 1. Participants of the Swiss Paediatric Airway Cohort (SPAC) who were included in the diagnostic performance analysis of fractional exhaled nitric oxide (FeNO)

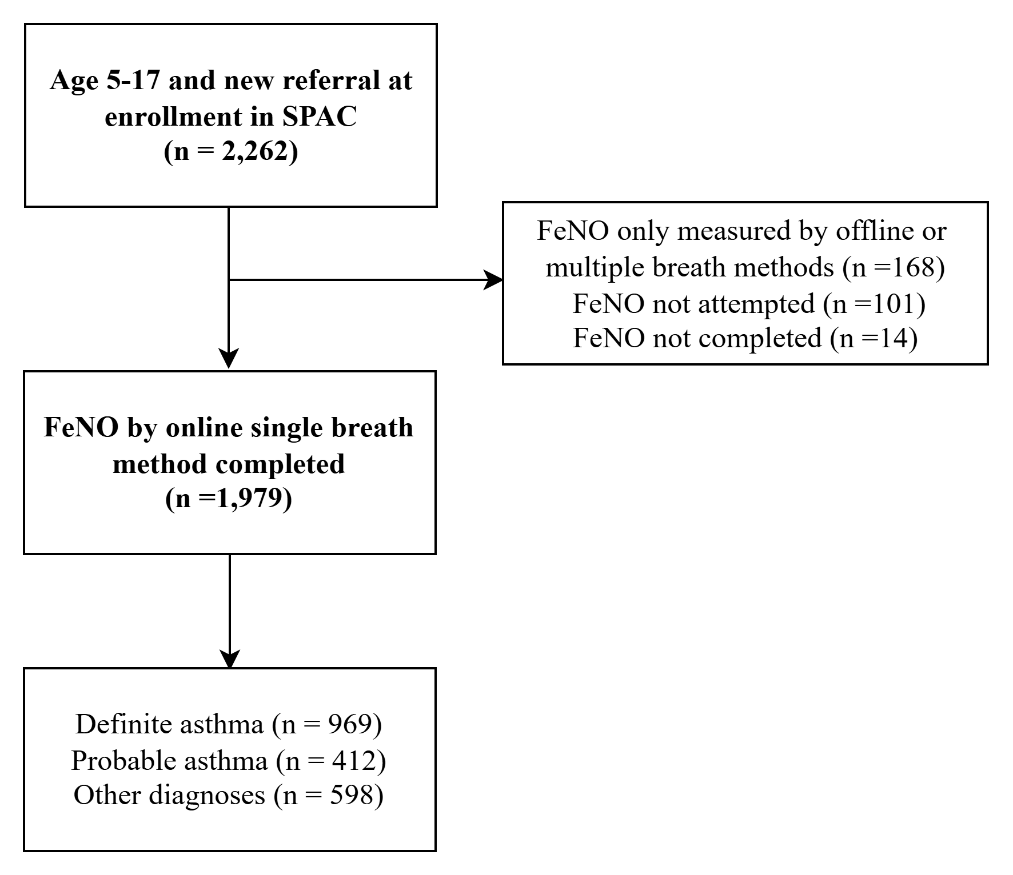

Abbreviations, FeNO: fractional exhaled nitric oxide, SPAC: Swiss Paediatric Airway Cohort

Supplementary table 1. Frequency of fractional exhaled nitric oxide (FeNO) measurement across the centres participating in the Swiss Paediatric Airway Cohort

|  | **Online single breath measured** | **Offline/online multiple breath only** | **FeNO not measured** |
| --- | --- | --- | --- |
| **Total (N = 2,262)** | 1,979 (87%) | 168 (7%) | 115 (5%) |
| **SPAC centre** |  |  |  |
| A (N = 710) | 610 (86%) | 59 (8%) | 41 (6%) |
| B (N = 402) | 327 (81%) | 58 (14%) | 17 (4%) |
| C (N = 346) | 337 (97%) | 0 (0%) | 9 (3%) |
| D (N = 269) | 238 (88%) | 23 (9%) | 8 (3%) |
| E (N = 223) | 182 (82%) | 28 (13%) | 13 (6%) |
| F (N = 118) | 111 (94%) | 0 (0%) | 7 (6%) |
| G (N = 96) | 88 (92%) | 0 (0%) | 8 (8%) |
| H (N = 51) | 47 (92%) | 0 (0%) | 4 (8%) |
| I (N = 47) | 39 (83%) | 0 (0%) | 8 (17%) |

Abbreviations, FeNO: fractional exhaled nitric oxide, SPAC: Swiss Paediatric Airway Cohort

|  | **Overall**  **N = 2,262** | **FeNO measurement** | |  |
| --- | --- | --- | --- | --- |
| **Characteristics** |  | **Online single breath measured**  **N = 1,979** | **Other methods or not measured**  **N = 283** | **p-value** |
| Age in years, median (IQR) | 9.0 (7.0, 12.0) | 10.0 (7.0, 12.0) | 6.0 (5.0, 8.0) | <0.001 |
| Male | 1,279 (57%) | 1,104 (56%) | 175 (62%) | 0.055 |
| BMI z-score, median (IQR) | 0.21 (-0.44, 1.00) | 0.22 (-0.43, 1.02) | 0.20 (-0.51, 0.92) | 0.28 |
| Self-reported symptoms and events in the last 12 months |  |  |  |  |
| Any wheeze | 1,669 (74%) | 1,469 (74%) | 200 (71%) | 0.2 |
| More than three attacks | 594 (26%) | 514 (26%) | 80 (28%) | 0.41 |
| Wheeze triggered by cold | 1,202 (53%) | 1,040 (53%) | 162 (57%) | 0.14 |
| Wheeze triggered by exercise | 1,263 (56%) | 1,141 (58%) | 122 (43%) | <0.001 |
| Wheeze triggered by pollen | 588 (26%) | 527 (27%) | 61 (22%) | 0.069 |
| Wheeze triggered by dust | 315 (14%) | 282 (14%) | 33 (12%) | 0.24 |
| Wheeze triggered by pets | 266 (12%) | 242 (12%) | 24 (9%) | 0.067 |
| Awakening due to wheeze | 698 (31%) | 597 (30%) | 101 (36%) | 0.06 |
| Dyspnoea due to wheeze | 972 (43%) | 855 (43%) | 117 (41%) | 0.55 |
| Emergency visit due to  respiratory symptoms | 858 (38%) | 729 (37%) | 129 (46%) | 0.005 |
| Hospitalisations due to  respiratory symptoms | 186 (8%) | 153 (8%) | 33 (12%) | 0.024 |
| Cough more than others | 1,107 (49%) | 945 (48%) | 162 (57%) | 0.003 |
| Self-reported allergic rhinitis | 910 (40%) | 810 (41%) | 100 (35%) | 0.073 |
| Asthma diagnosis | 1,566 (69%) | 1,381 (70%) | 185 (65%) | 0.13 |

Supplementary table 2. Clinical characteristics of children with and without fractional exhaled nitric oxide (FeNO) measured by online single breath method in the Swiss Paediatric Airway Cohort

Abbreviations, FeNO: fractional exhaled nitric oxide, IQR: interquartile range, p-value was derived by Pearson’s Chi-squared test or Wilcoxon rank sum test.

Supplementary table 3. Main diagnosis of children who were not diagnosed with asthma in the Swiss Paediatric Airway Cohort (n = 598)

| **Final main diagnosis** | **n (%)** |
| --- | --- |
| Exercise-related diagnoses |  |
| Extra-thoracic dysfunctional breathing | 92 (15%) |
| Thoracic dysfunctional breathing | 89 (15%) |
| Exercise-induced symptoms of unknown aetiology | 65 (11%) |
| Cough-related diagnoses |  |
| Upper airway cough | 55 (9%) |
| Cough of unknown or other aetiology | 65 (11%) |
| Bronchial hypersensitivity (no clinical asthma) | 35 (6%) |
| Postinfectious cough | 21 (4%) |
| Habitual cough | 7 (1%) |
| Upper airway diagnoses |  |
| Allergic rhinitis/allergy related respiratory symptoms | 89 (15%) |
| Upper airway issues (e.g., snoring) | 13 (2%) |
| Chronic rhinosinusitis | 4 (1%) |
| Infection related diagnoses |  |
| Respiratory tract infections | 14 (2%) |
| Protracted bacterial bronchitis | 12 (2%) |
| Other diagnoses | 37 (6%) |

Supplementary table 4. Cut-offs of fractional exhaled nitric oxide (FeNO) required to achieve selected sensitivity and specificity levels in the Swiss Paediatric Airway Cohort

|  | **Target value (%)** | **Sensitivity (%)** | **FeNO cut-off (ppb)** | **Specificity (%)** | **FeNO cut-off (ppb)** |
| --- | --- | --- | --- | --- | --- |
| **Sensitivity** | 80 |  |  | 31 | 8.8 |
|  | 90 |  |  | 14 | 6 |
|  | 95 |  |  | 6 | 4.5 |
| **Specificity** | 80 | 48 | 21.3 |  |  |
|  | 90 | 32 | 33.6 |  |  |
|  | 95 | 24 | 44 |  |  |

Abbreviations, FeNO: fractional exhaled nitric oxide, ppb: parts per billion

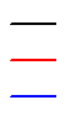
Supplementary figure 2. Diagnostic performance of fractional exhaled nitric oxide (FeNO) in children with and without inhaled corticosteroid (ICS) use (only among children with available information from the medical record, n = 448) in the Swiss Paediatric Airway Cohort

With ICS use

Without ICS use

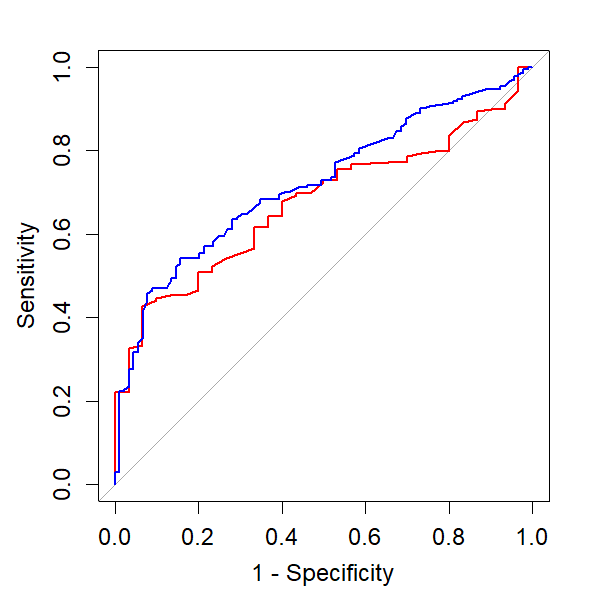

| **Cutoff (ppb)** | **Sensitivity** | **Specificity** | **PPV** | **NPV** | **Youden's Index** |
| --- | --- | --- | --- | --- | --- |
| **With ICS use (n = 189, asthma 84%, AUC 0.67, 95%CI 0.59-0.76, p < 0.001)** | | | | | |
| ≥22 | 44 (37–52) | 90 (74–97) | 96 (89–99) | 23 (17–32) | 0.34 |
| **≥23** | 42 (35–50) | 93 (79–98) | 97 (90–99) | 23 (17–32) | 0.36 |
| ≥25 | 40 (33–48) | 93 (79–98) | 97 (90–99) | 23 (16–31) | 0.34 |
| ≥35 | 27 (21–34) | 97 (83–99) | 98 (88–100) | 20 (14–27) | 0.24 |
| **Without ICS use (n = 259, asthma 66%, AUC 0.72, 95%CI 0.66-0.78, p < 0.001)** | | | | | |
| ≥22 | 55 (48–63) | 80 (70–87) | 84 (76–90) | 48 (40–56) | 0.35 |
| **≥23** | 54 (47–61) | 84 (75–90) | 87 (79–92) | 49 (41–57) | 0.38 |
| ≥25 | 49 (41–56) | 87 (78–92) | 87 (79–93) | 47 (39–55) | 0.35 |
| ≥35 | 35 (29–43) | 93 (86–97) | 91 (82–96) | 43 (36–50) | 0.29 |

Data are presented as n or n (95%CI), values with the cutoff with the highest Youden’s index within each subgroup (bold), optimal cutoff among all children in SPAC (22 ppb), ERS cutoff (25 ppb) and BTS/NICE/SIGN cutoff (35 ppb). P- values were obtained by testing each AUC against 0.5 using DeLong’s method. DeLong’s test for difference in the ROC for with ICS vs without ICS use: p = 0.39.

Abbreviations, AUC: area under the curve, ICS: inhaled corticosteroid, PPV: positive predictive index, NPV: negative predictive index, FeNO: fractional exhaled nitric oxide, ppb: parts per billion

Supplementary figure 3. Diagnostic performance of fractional exhaled nitric oxide (FeNO) for asthma in children in the Swiss Paediatric Airway Cohort using alternative reference standards

| Cutoff (ppb) | Sensitivity | Specificity | PPV | NPV | Youden's Index |
| --- | --- | --- | --- | --- | --- |
| **Definite vs probable and not asthma (n = 1,979, asthma 49%, AUC 0.66, 95%CI 0.63-0.68, p < 0.001)** | | | | | |
| **≥22** | 53 (49–56) | 74 (71–77) | 66 (62–69) | 62 (59–65) | 0.27 |
| ≥25 | 48 (44–51) | 78 (75–80) | 67 (64–71) | 61 (58–63) | 0.25 |
| ≥35 | 36 (33–39) | 86 (84–88) | 71 (67–75) | 58 (56–61) | 0.22 |
| **ICS use at 1-2y vs no use^#^ (n = 1,213, ICS use 53%, AUC 0.58, 0.55-0.62, p < 0.001)** | | | | | |
| **≥20** | 51 (47–55) | 64 (60–68) | 62 (57–66) | 54 (50–57) | 0.15 |
| ≥22 | 48 (44–52) | 67 (63–71) | 62 (58–67) | 53 (49–57) | 0.15 |
| ≥25 | 42 (39–46) | 73 (69–76) | 64 (59–68) | 53 (49–56) | 0.15 |
| ≥35 | 31 (27–35) | 82 (78–85) | 66 (60–71) | 51 (48–54) | 0.13 |

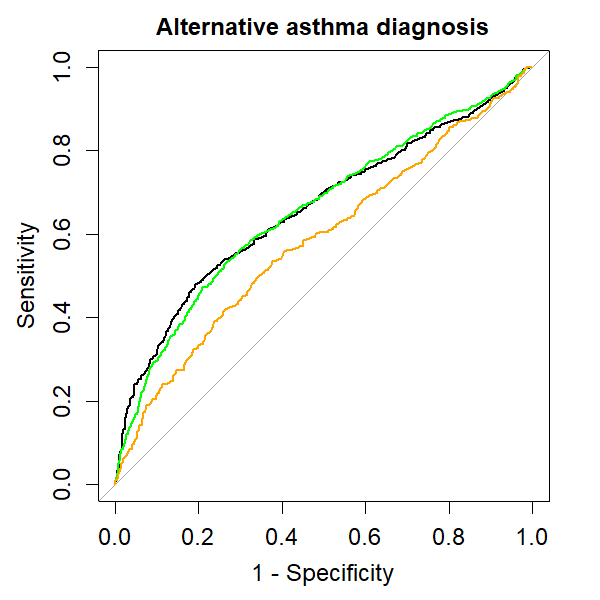

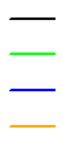

Main reference standard: Definite + probable vs not asthma

Definite vs probable asthma + not asthma

Asthma controller use 1 year later (as a proxy of diagnosis)

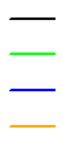

Data are presented as n or n (95%CI), values with the cutoff with the highest Youden’s index for each reference standard (bold), optimal cutoff in primary analysis (22 ppb), ERS cutoff (25 ppb) and BTS/NICE/SIGN cutoff (35 ppb). P- values were obtained by testing each AUC against 0.5 using DeLong’s method.

^#^ICS use includes the use of ICS or ICS-LABA, at any duration reported in the follow up questionnaire at 1y or 2y (when the 1y questionnaire is unavailable).

Abbreviations, AUC: area under the curve, ICS: inhaled cortico-steroid, ICS-LABA: Inhaled corticosteroids/long-acting beta2-agonists, TP: true positive, FP: false positive, FN: false negative, TN: true negative, PPV: positive predictive index, NPV: negative predictive index, FeNO: fractional exhaled nitric oxide, ppb: parts per billion

Reference

1.	de Onis M, Onyango AW, Borghi E, Siyam A, Nishida C, Siekmann J. Development of a WHO growth reference for school-aged children and adolescents. Bull World Health Organ. 2007;85(9):660-7.

2.	Whiting P, Rutjes AW, Reitsma JB, Glas AS, Bossuyt PM, Kleijnen J. Sources of variation and bias in studies of diagnostic accuracy: a systematic review. Ann Intern Med. 2004;140(3):189-202.
